# Understanding the relationship between the presence of vegetation and the spread of canine visceral leishmaniasis in Camaçari, Bahia State, Northeastern Brazil

**DOI:** 10.1101/2023.08.31.23294879

**Authors:** Freya N. Clark, Manuela da Silva Solcà, Deborah Bittencourt Mothé Fraga, Claudia Ida Brodskyn, Emanuele Giorgi

## Abstract

Leishmaniasis is a vector-borne disease spread by female phlebotomine sandflies (*Lutzomyia longipalpis*). The most severe form of the disease is visceral leishmaniasis (VL), which can cause fever, hepatosplenomegaly, weight loss and pancytopenia. Domestic canines are the main reservoir for human cases in Brazil because they live in close proximity and can remain asymptomatic for long periods of time. Consequently, sole treatment of human cases will not contain the spread of the disease. Current methods of control have been unsuccessful, and thus a better understanding of the canine transmission and the effect of their environment is required. Vegetation is one of the main risk factors for VL that affects the distribution of phlebotomine sandflies. Using geostatistical models, we aim to further understand the effect of vegetation on canine VL in the community of Camaçari, northeastern Brazil. The risk due to vegetation is quantified using the average of the normalised vegetation index (NDVI) for all pixels within each dog’s home range. We found that an increase in NDVI of 0.1 led to an 1.21-fold increase in the odds of canine visceral leishmaniasis, on average, suggesting that coastal vegetation has a particularly strong correlation with VL.

**Author summary:** Leishmaniasis is a disease spread by female phlebotomine sandflies when feeding from mammal blood. Visceral leishmaniasis (VL), the most severe form of the disease, causes fever, weight loss and swelling of internal organs. The vast majority of human VL cases in the Americas occur in Brazil, where domestic canines act as disease pools due to their close proximity to humans and high proportion of asymptomatic cases. Due to a lack of testing and reporting of canine VL cases, authorities have been unable to control transmission. Understanding how VL spreads to canines is imperative to develop new prevention and control strategies. Phlebotomine sandflies feed on plant sap and nectar, and lay eggs around tree roots, hence, we suspect that a mammal would be more likely to contract VL when living near vegetation. We investigated how a dog’s proximity to vegetation affects its chances of contracting VL. We used a geostatistical model that combined the measure of the vegetation with the spatial correlation of the sampled locations of the dogs. Our model estimated that, on average, an increase of 0.1 in the measure of vegetation led to an 1.21-fold increase in the odds that a canine contracted VL.

## Introduction

Leishmaniasis is one of the world’s most neglected tropical diseases [1]. The most severe form is visceral leishmaniasis (VL), which attacks the internal organs of the host, causing swelling of the spleen, liver and lymph nodes [2–4]. Female phlebotomine sandflies (*Lutzomyia longipalpis*) transmit the disease when feeding on mammal blood [4, 5]. Over 97% of human VL cases in the Americas are within Brazil, and human vaccinations are not currently available [3, 6]. Since nearly 50% of infected domestic canines (*Canis familiaris*) are asymptomatic and can remain asymptomatic for several years, they have become the main reservoir for human infection in Brazil [3, 5, 7]. Additionally, symptomatic dogs exhibit a variety of different clinical signs, so laboratory analysis is required for diagnosis [7]. Due to the zoonotic transmission occurring in Brazil, solely preventing and treating human cases is not sufficient to prevent new cases; reducing the prevalence of leishmaniasis in dogs is imperative to prevent both human and canine cases.

The prevalence of human visceral leishmaniasis is well documented, and several studies have revealed that human VL cases in Brazil are intrinsically linked to cases of canine VL [8–11]. However, there is a substantial lack of testing and reporting of canine cases [12]. Canine vaccinations are available, however these have a relatively low efficacy (68%-71%) [7]. Medicated collars and spot-ons have a higher efficacy of 90% under laboratory conditions [13], however use is limited since most owners of at risk dogs cannot afford these treatments, and many dogs in Brazil are stray [2]. Dog culling is a popular method of attempted disease control in Central and South America [14]. However, this has been proven ineffective for multiple reasons: low sensitivity and specificity of diagnostic tests, culled dogs are often replaced by younger dogs who are more at risk of disease, widespread use is not financially viable, and the practice is subject to ethical criticism [2, 14]. The use of insecticides has proven effective in reducing contact between sandflies and both humans and dogs [14]. However, this practice poses a risk to non-target organisms and visceral leishmaniasis is still widespread in Brazil so a deeper understanding of how dogs are contracting VL is necessary to advance prevention strategies.

In this study, we focus on the municipality of Camaçari in Bahia, Northeastern Brazil. Vegetation is an important factor that affects the sandfly population in several ways: plant sap and nectar are their main food source of sandflies; they tend to lay eggs around tree roots and animal burrows; once hatched, the larvae feed on decaying vegetation and animal faeces [2, 13]. Information on vegetation can thus be used as a proxy for the sandfly population, which can be difficult to estimate, and aid the mapping of VL risk in a region of interest, allowing authorities to target prevention strategies on areas at highest risk [15].

Previous studies present conflicting results with regards to the relationship between sandflies and vegetation. A number of studies suggest that vegetation is inversely correlated to VL cases, since VL has an urban transmission cycle [16–18]. On the other hand, increased vegetation growth, such as during periods of high rainfall, have been found to cause an influx in sandfly population [19–22]. This suggests that the connection between vegetation and VL is complex, and requires further investigation.

Several studies have used geostatistical modelling to map VL risk among humans in Brazil. For example, Werneck and Maguire [23] analysed the incidence of human VL in Piauí. The data was aggregated by consolidated census tract, and they found that living in areas covered by green vegetation was positively associated with the incidence of VL. In another study, Karagiannis-Voules *et al*. [24] carried out a country-wide analysis and found that the incidence of human VL was not associated with the presence of vegetation at municipality level. However, these studies use aggregated data which does not allow them to obtain an unbiased estimate of the effect of vegetation, as a result of the ecological fallacy. To the best of our knowledge there are no previous studies that have carried out a geostatistical analysis canine VL cases and how these are affected by vegetation.

To address this knowledge age, this study aims to model the relationship between the presence of vegetation around a household and the occurrence of visceral leishmaniasis in dogs using model-based geostatistical methods of analysis [15]. Geostatistical models have been extensively used to account for the residual spatial variation in disease risk and enable mapping of health outcomes throughout an area of interest [15]. Accounting for spatial correlation also allows us to obtain reliable estimates of uncertainty in the regression relationships which is essential for the primary objective of our study.

## Materials and methods

### Study area

Data regarding canine cases of visceral leishmaniasis in the municipality of Camaçari in Bahia, Northeastern Brazil, was obtained from the Federal University of Bahia (UFBA). Camaçari is 785 km^2^ and contains three districts: Sede de Camaçari, Abrantes, and Monte Gordo [25]. Dogs were sampled from all three districts from May 2011 to June 2012. Estimates of the canine population were obtained from the Camaçari Center for Disease Control, and 20% of the dog population was randomly sampled from each neighbourhood in the entire municipality. Dogs living in the same household were given the same geographical location. A map of the study area is shown in Fig 1. As Brazil is a low-to-middle-income country, disease registries are not a viable option for epidemiological studies [15, 26]. Therefore, household surveys were conducted to collect the data. Demographic information for Camaçari was obtained from the 2010 census.

**Fig 1.**
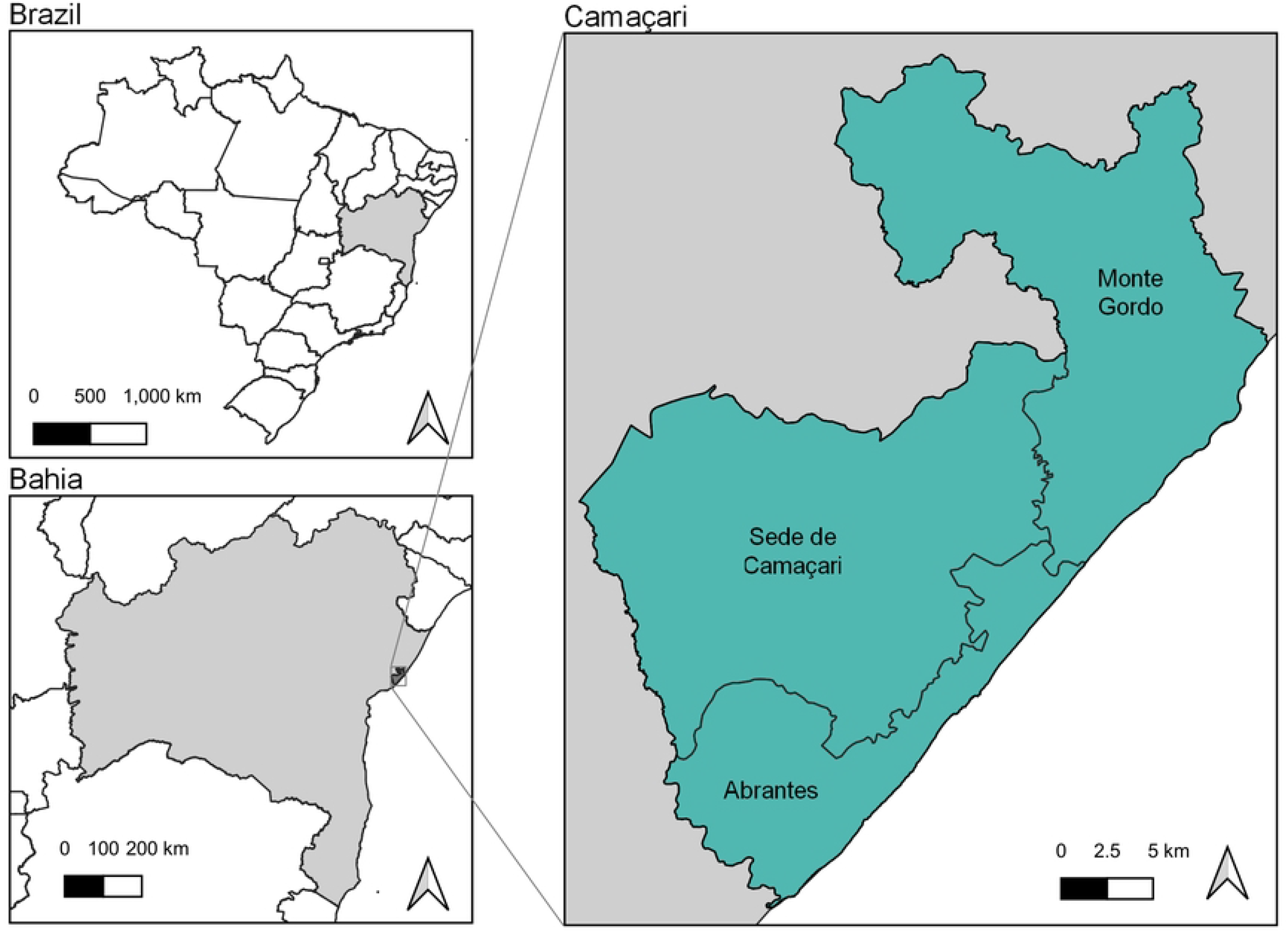
Camaçari geography. Map showing the location of Camaçari in Bahia, Brazil.

### Dataset

Information on 800 dogs in 530 households was collected during the study. The location and test results for each dog were combined with the household questionnaire and results of a physical examination of the dog. We consider the canine VL diagnosis to be the binary outcome for our study; a total of 21 dogs were excluded from analysis due to a missing canine VL diagnosis. Additionally, 2 dogs were removed due to incorrect location information, leaving 777 dogs in 520 households. Indicators of whether the dog lives on the coast and whether there were other infected dogs in the household were added, since these were identified as important factors in the literature. The area defined as coastal, and the geographical locations of dogs included in the study are shown on a map of Camaçari in Fig 2. The majority of the sampled dogs live either in the capital city, Sede de Camaçari, or on the coast of Abrantes and Monte Gordo.

**Fig 2.**
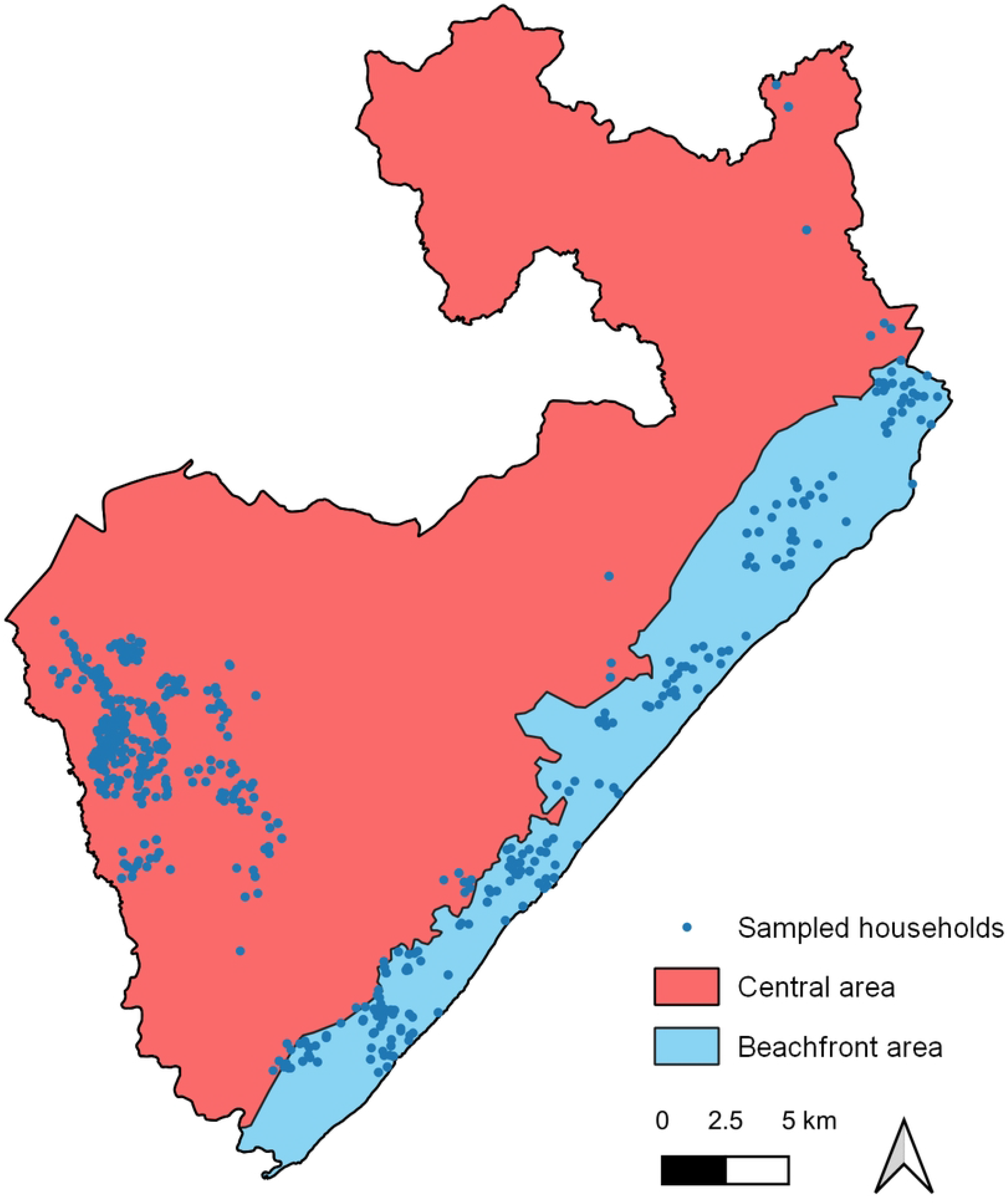
Dataset locations. Map of Camaçari showing the area assigned as coastal (light blue) and the locations of the households involved in the study (dark blue).

#### Ethics statement

All experiments involving animals were performed in accordance with the institutional review board of the Gonçalo Moniz Institute (IGM - Fiocruz - Bahia/Brazil), and were approved by the CEUA of Fiocruz - Bahia (protocol CEUA 017/2010 and 007/2013).

#### Diagnosis

Canine visceral leishmaniasis was diagnosed by collecting splenic aspirate, skin biopsy, and whole blood samples every 6 months [27]. Chromatographic immunoassay and ELISA confirmation assay were used to detect antibodies, in line with the Brazilian Ministry of Health standards [27, 28]. Dogs were considered infected for the whole study period if at least one tissue was positive for Leishmania infantum infection at any time throughout the study period [27]. Dogs were assessed by veterinarians regarding the presence of clinical manifestation suggestive of leishmaniasis.

### Vegetation index

#### Satellite imagery

In order to map out the vegetation in Camaçari, Landsat 7 satellite imagery was obtained from the U.S. Geological Survey (USGS). These images have a resolution of 30 m *×* 30 m. During the study period, the majority of available satellite images are mostly or completely obscured by cloud cover, preventing vegetation indexing in large parts of the study area. Additionally, in 2003 there was a permanent failure in the Scan Line Corrector which results in 22% of the pixels in Landsat 7 images being lost [29]. Therefore, a combination of 11 Landsat 7 satellite images taken during the 2011 and 2012 were used to create a complete image of the landscape of Camaçari. Clouds and large shadows were removed from the satellite images in QGIS with the aid of the CloudMasking plugin v22.2.25. Large bodies of water were also removed, as they produce a very low vegetation value, and proximity to water is not of interest in this study. Fig 3 shows the combined RGB image for Camaçari with large bodies of water removed.

**Fig 3.**
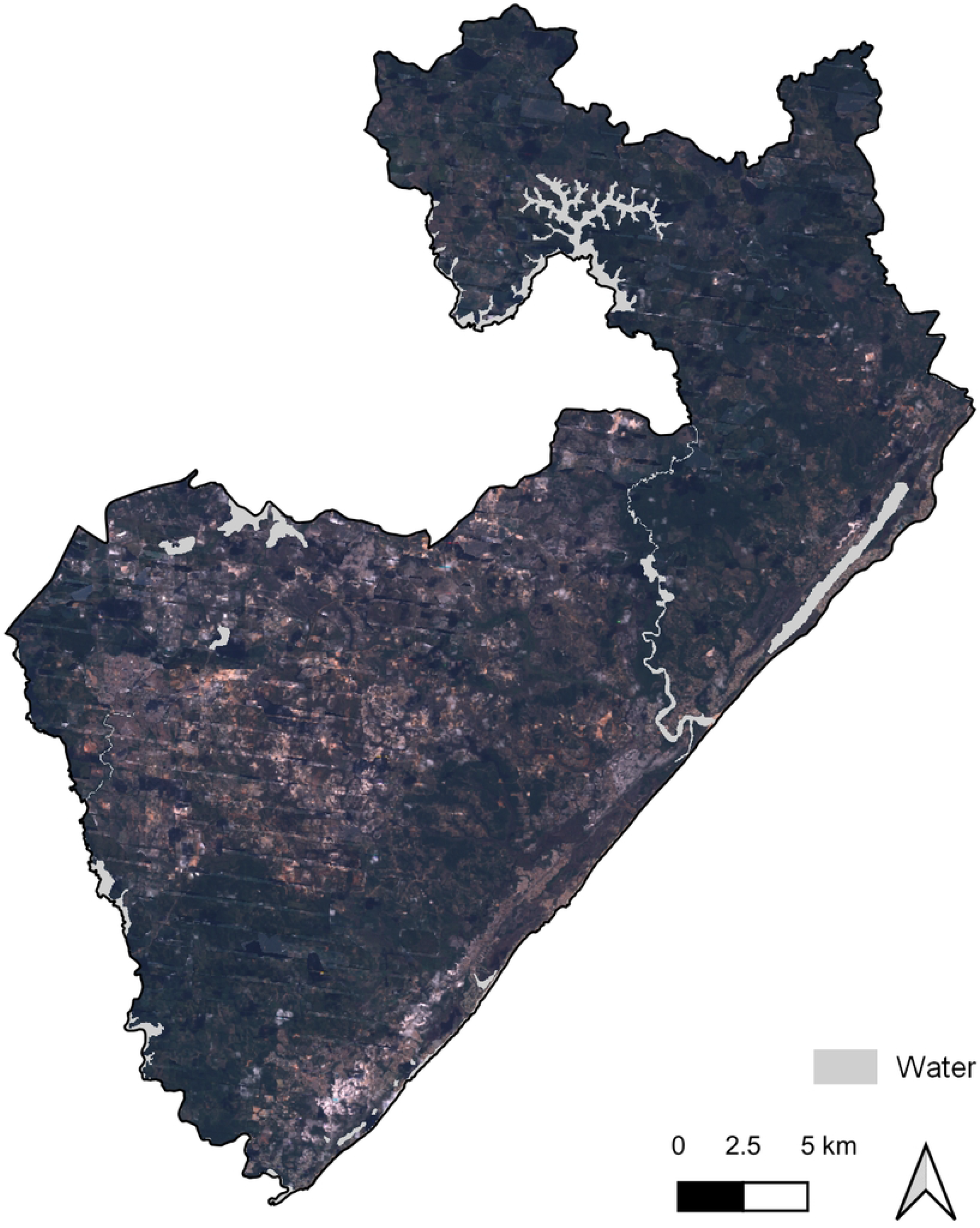
Satellite image of Camaçari. Combination of Landsat 7 satellite images of Camaçari.

#### Vegetation index calculation

Vegetation Index calculations were performed in Quantum Geographic Information System (QGIS) v3.22.7. For this analysis, the Normalized Difference Vegetation Index (NDVI) and Enhanced Vegetation Index (EVI) were both considered as methods of quantifying the vegetation cover. Since the majority of the study area is not covered by dense vegetation, we did not find an appreciable difference between the two measures. Hence, we proceed with NDVI, which is the simpler of the two calculations. The NDVI calculates the averaged difference between the near infrared and red bands of light, giving a value between -1.0 and +1.0 for each pixel in the satellite image [30]. A low vegetation index refers to rocks, sand, or snow, and a high vegetation index refers to areas of dense vegetation [31]. Fig 4 shows the calculated NDVI values for Camacari.

**Fig 4.**
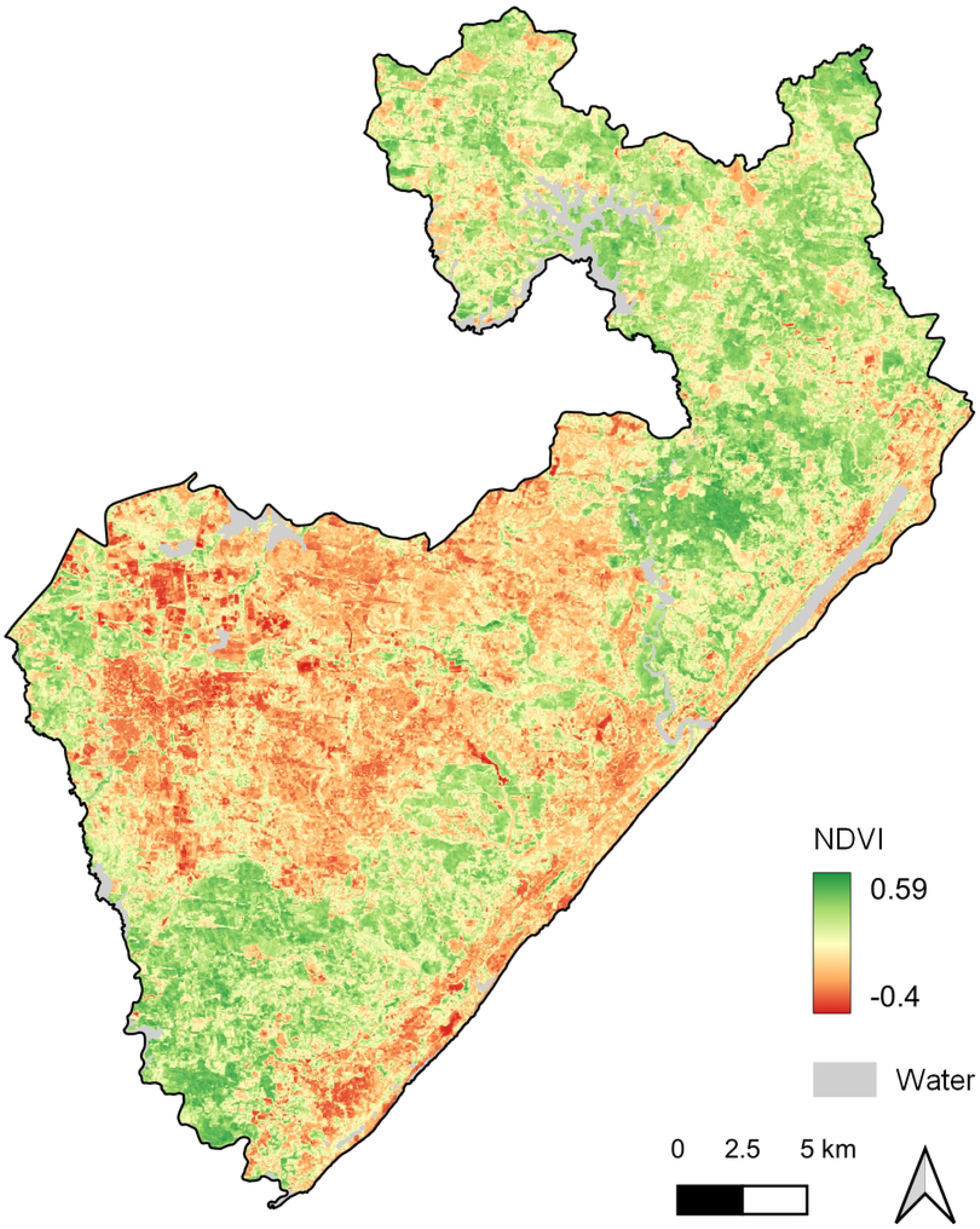
NDVI of Camaçari. Map of the calculated NDVI values for Camaçari.

#### Canine home range

In order to assign a Vegetation Index to each dog in the study, the area which they frequently travel around their home must be determined; this is referred to as their home range [32]. There is little information about where and how far free-roaming domestic canines travel, however it is estimated that they can cover on average 65 ha [32–35]. This gives a radius of approximately 455 m. A distance matrix was produced between each canine in the study and the coordinates of the pixels in the satellite image. Any pixels which fall inside the radius were averaged to give each canine’s NDVI value.

### Geostatistical modelling

For the *j*^*th*^ sampled dog, let *Y*_*j*_(*x*_*i*_) : *i* = 1, …, *n* be the outcome of the test for canine visceral leishmaniasis, and *x*_*i*_ be the discrete set of locations of the dogs’ households within Camaçari. Asymptomatic dogs have been found to infect sandflies, so it is suggested that symptomatic and asymptomatic dogs should be considered equally [1, 7]. Since 50% of the sampled dogs were the only dog included in their household, each dog is considered individually for statistical analysis, so *Y*_*j*_ is a Bernoulli random variable, where *Y*_*j*_(*x*_*i*_) = 1 or 0 corresponding to a positive or negative result, respectively. Hence, we write Prob(*Y* = 1) = *π*. To allow for spatial variation in the cases, we allowed *π* to vary according to both measured location-specific covariates (*μ*(*x*)) and unexplained residual spatial variation (*S*(*x*)), as follows,

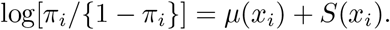

Here, the mean function *μ*(*x*) is a linear regression,

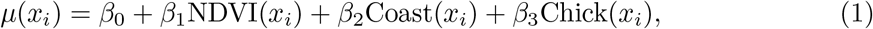

where NDVI(*x*_*i*_) is the normalised vegetation index (NDVI) at location *x*_*i*_, Coast(*x*_*i*_) is a binary indicator for whether location *x*_*i*_ is on the coast of Camaçari, Chick(*x*_*i*_) is a binary indicator for whether chickens are kept at location *x*_*i*_, and *β* are the regression parameters. We defined *S*(*x*) to be a zero-mean stationary Gaussian process.

The rationale for the chosen covariates is as follows. The coastal areas of Abrantes and Monte Gordo are less built up than the city of Camaçari, with more vegetation present, and more sandflies are present along the coast. Therefore, dogs living on the coast are more likely to come into contact with sandflies and contract VL. Sandflies are often found living in chicken coops since they provide shelter from winds, animals to feed from and faeces to lay eggs in [11, 36], so any dogs living in a household which keeps chickens are more likely to come into contact with sandflies, and hence VL. Family farming of livestock such as chickens is highly present in rural areas of northeastern Brazil, where vegetation is more abundant [37]. A directed acyclic graph showing these relationships is presented in the supplementary material(8).

The following covariates were also considered, but did not result in improved performance: the socio-economic status of the household; the size and age of the dog; whether the dog roams outdoors freely; whether prevention measures were used; the presence of open sewage.

We estimated the model parameters of Eq (1) using the Monte Carlo maximum-likelihood (MCML) method implemented in the PrevMap package [38]. Laplace estimation does not perform well for binary outcomes [39], but was used to approximate initial values to input into the MCML model for faster convergence.

Validation of the MCML model was carried out using a variogram-based procedure, which tested the compatibility of the adopted spatial structure with the data. Hierarchical clustering was used due to its sensitivity to outliers. The procedure was carried out as follows: Step 1. 1000 data sets were simulated under the fitted model in Eq (1); Step 2. A variogram was computed for each simulated data set, using the residuals from a the model in Eq (1) with *S*(*x*) = 0 for all locations *x*; Step 3. A 95% confidence interval was computed using the resulting 1000 variograms; Step 4. A variogram was computed as before except the original data was used instead of the simulated data. This is defined as the observed variogram. If the observed variogram fell within the 95% confidence interval, the spatial correlation for the model in Eq (1) was compatible with the canine VL data.

Additionally, the model in Eq (1) was compared with an model which does not use vegetation as a covariate to identify the effect of including vegetation. The mean function for this model was defined as

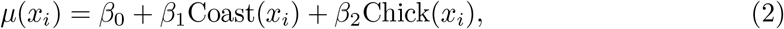

From the GLGM parameter estimates, we used spatial prediction to find the odds surface over Camaçari, denoted *A*. The predictive target *T*^*^ was defined as

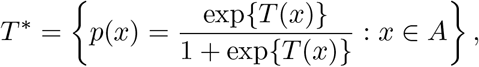

where *T* (*x*) is the model given in Eq (1). We approximated the area of interest *A* with a regular grid 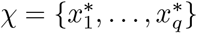 of *q* = 4996 precise locations in Camaçari. The predictive target *T*^*^ was be calculated from the fitted values of the GLGM model and the spatial surface. This allowed us to create a map of the odds surface and the find the probability that the odds exceeded 1 at each location on the grid. The uncertainty of the estimates were determined by mapping a quantile surface for the 2.5% and 97.5% quantiles. Further details of the Laplace and MCML method, and calculation of the predictive target can be found in Diggle and Giorgi’s work [15].

## Results

The presence of spatial clustering in the data was confirmed by the Global Moran’s index, with a *p*-value *<* 0.001. The parameter estimates and confidence intervals (CI) for our fitted geostatistical model using the Monte Carlo method are shown in Table 1, where NDVI was used as the measure of vegetation.

**Table 1.** MCML model parameter estimates. Maximum likelihood estimates and confidence intervals (CI) for the odd ratios of the parameters in the models described in Eq (1) and Eq (2). Significant coefficients are indicated with *.

| Parameter | Model (1) |  | Model (2) |  |
| --- | --- | --- | --- | --- |
|  | Estimate | 95% CI | Estimate | 95% CI |
| Intercept | 0.151 | (0.103, 0.219)* | 0.174 | (0.126, 0.242)* |
| Vegetation | 12.109 | (1.258, 116.545)* | - | - |
| Live On Coast | 2.608 | (1.628, 4.176)* | 3.190 | (2.105, 4.835)* |
| Keep Chickens | 1.215 | (0.773, 1.910) | 1.347 | (0.905, 2.004)* |
| Variance ($\sigma^2$) | 1.895 | (1.410, 2.546)* | 2.114 | (1.620, 2.759)* |
| Scale ($\phi$) | 0.871 | (0.605, 1.252) | 0.922 | (0.683, 1.246) |

The scaling parameter *ϕ* indicates the distance that the spatial correlation persists through the exponential correlation function as follows: 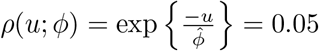. Hence, the spatial correlation *ρ*(*u*; *ϕ*) decays to 0.05 at 2.61km.

The empirical semi-variogram fell within the 95% tolerance intervals, so we concluded that the MCML model with compatible with the data. A plot of the empirical semi-variogram is presented in the supplementary material(9).

As shown in Table 1, the inclusion of vegetation in Model (1) reduced the amount of residual spatial variation by 10%. Additionally, the average standard errors of the prediction grid were 1.519 and 1.784 for the Model (1) and Model (2), respectively. Therefore, vegetation contributed to explain the residual spatial variation by about 15%.

Fig 5 shows a comparison of the predictive map of the odds estimated by the MCML model for the case where no owners keep chickens, and the probability that the odds exceeds 1. The predicted odds range from 0.206 to 2.590, with the lowest odds in the capital city of Camaçari (west) and the high odds on the coast of Monte Gordo (north east). The exceedance probabilities are lowest in the capital city, with the lowest being 0.022 and the highest exceedance probabilities, with a maximum of 0.505, are found on the coast of Monte Gordo. The 2.5% and 97.5% quantiles for the MCMC model are shown in Fig 6.

**Fig 5.**
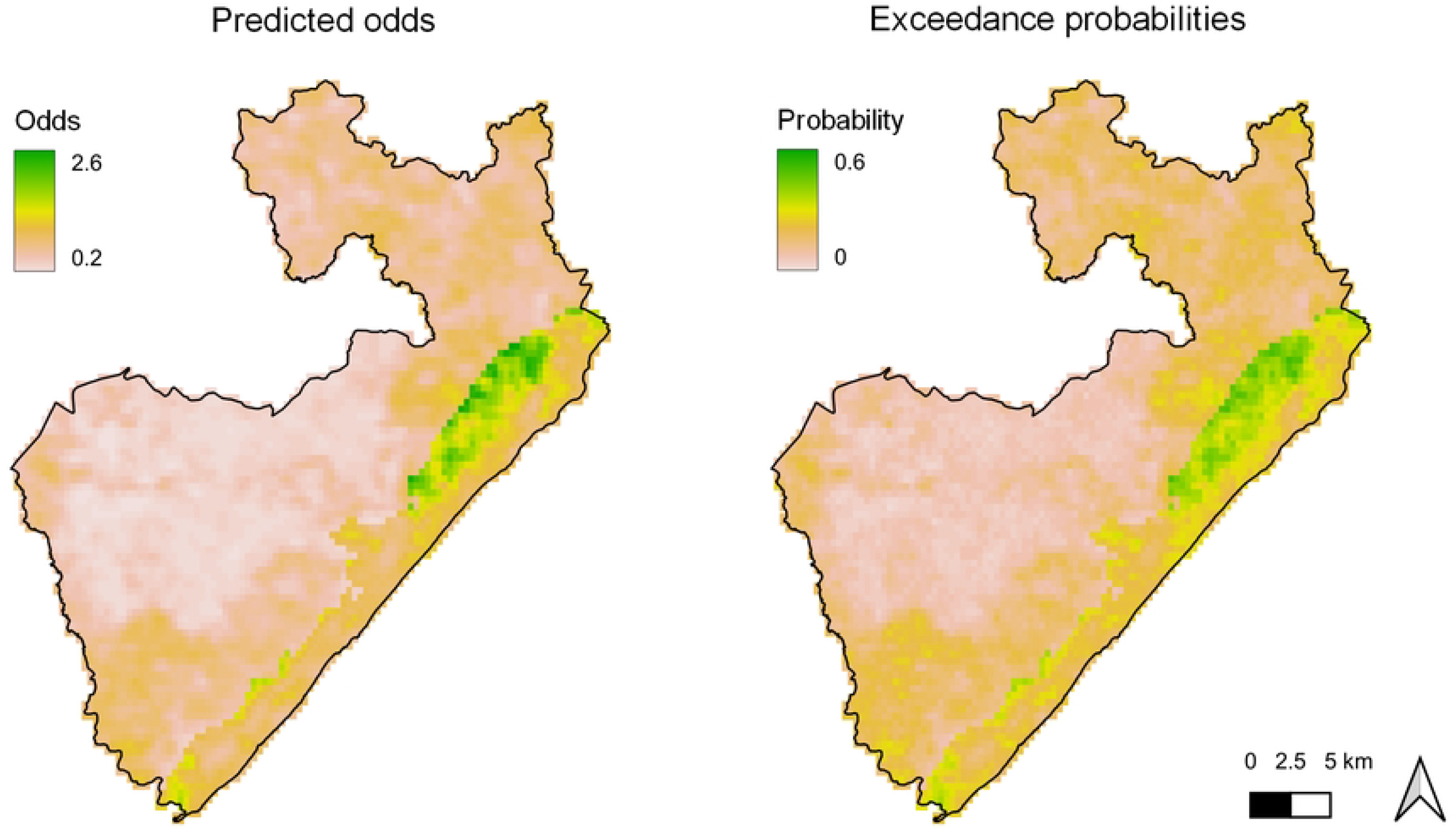
Predicted odds and exceedance probability. Maps of the predicted odds (left panel) and probability of exceeding odds of 1 (right panel). Odds were predicted using the MCML model assuming all owners do not keep chickens.

**Fig 6.**
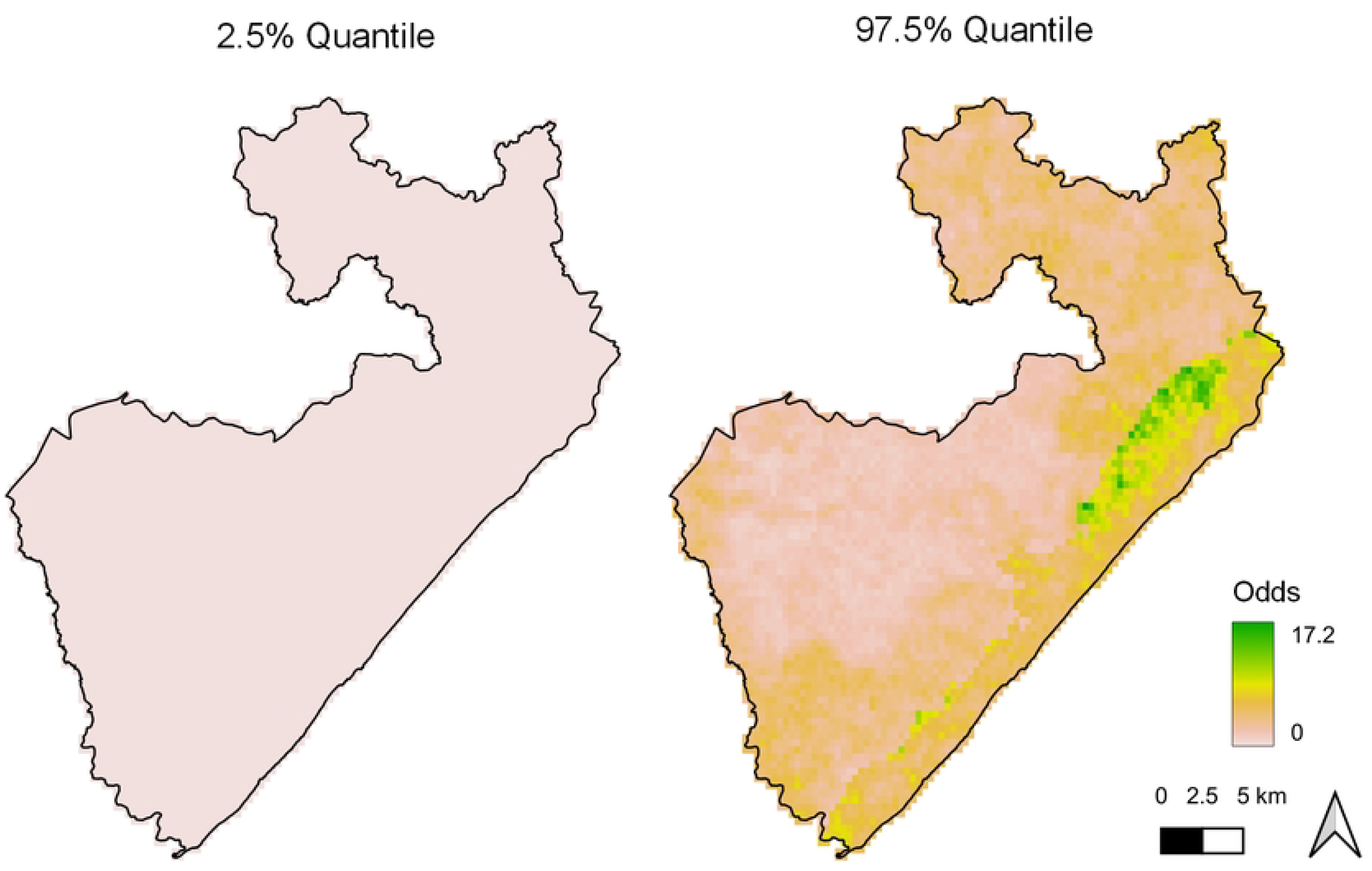
Predicted odds quantiles. Map of the 2.5% quantile (left panel) and 97.5% quantile (right panel) for the MCML model predicted odds.

Finally, Fig 7 shows a map of exp*{β*_1_NDVI(*x*_*i*_)*}*, the odds for the vegetation coefficient in the MCML model. The vegetation coefficient had the lowest impact on the odds in the capital city and along the coast of Camaçari. The most impact is found in the south of Abrantes and north of Monte Gordo.

**Fig 7.**
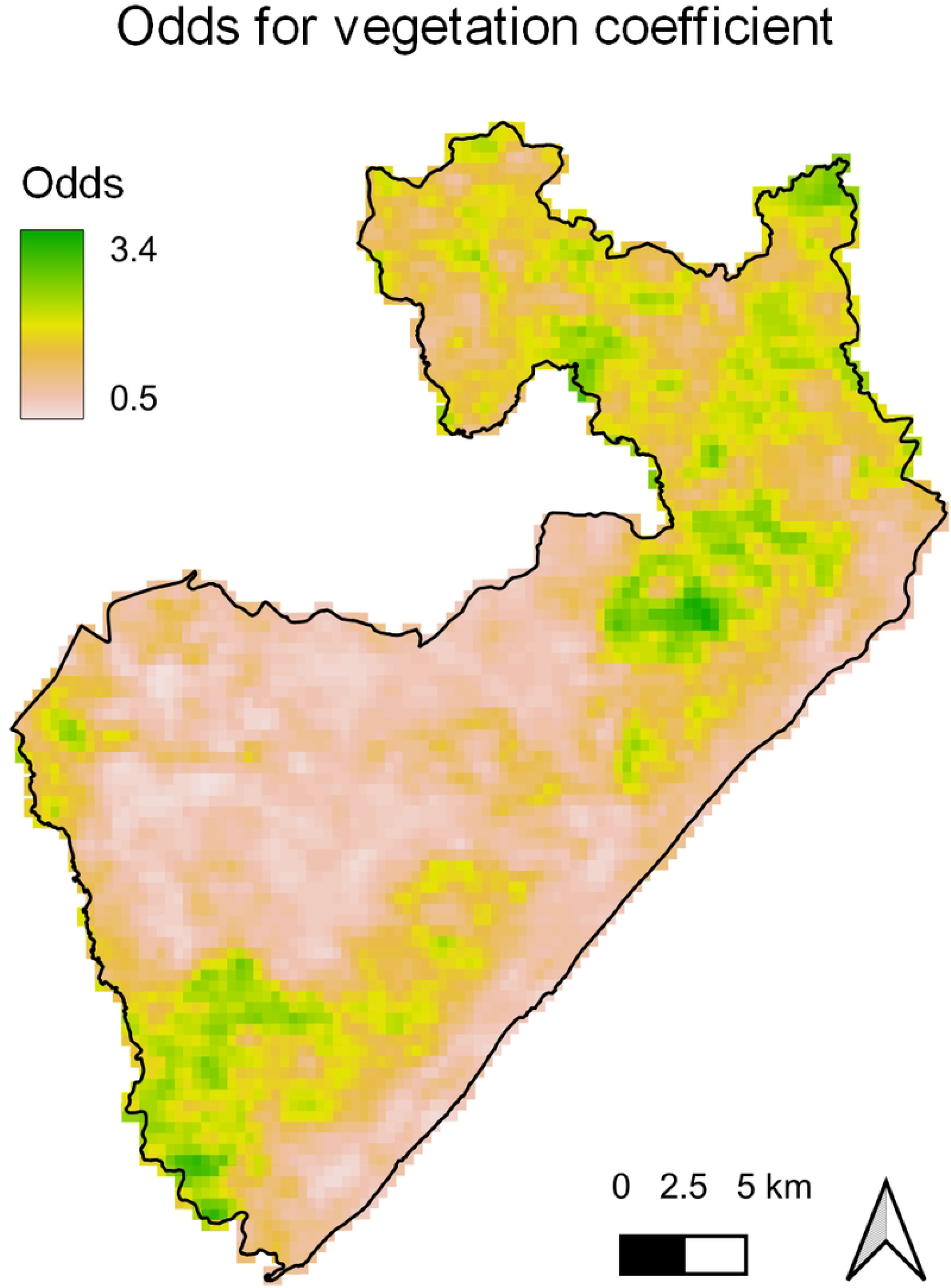
Predicted odds for vegetation coefficient. Map of the odds ratio for the vegetation coefficient of the MCML model.

**Fig 8.** Relationships between model variables. A directed acyclic graph showing the relationship between vegetation, canine visceral leishmaniasis and identified confounders. The arrow of interest is shown in bold.

**Fig 9.** Empirical semi-variogram. Empirical variograms (left panel) of the estimate residual variation (solid lines) and 95% tolerance bandwith (grey area). Null distribution (right panel) of the test statistic determining whether the variogram using the original data falls within the 95% confidence interval.

## Discussion

In this paper, we have applied geostatistical modelling to data collected from domestic canines in Camaçari, Brazil, to investigate the relationship between a dog’s proximity to vegetation and its odds of contracting canine visceral leishmaniasis. We used the normalised vegetation index (NDVI) as the measure of vegetation, and found that the highest risk of canine VL is found in the most vegetated parts of urban areas in Camaçari. The results have indicated that, on average, a 5% increase in vegetation led to an 1.21-fold increase in the odds of disease. In addition, we hypothesise that the type of vegetation present has a significant impact on this relationship, corresponding with the findings of Mota *et al* [40].

The estimates for Model (1) in Table 1 can be interpreted as follows: the intercept coefficient refers to the odds ratio of canine VL for a dog who does not live on the coast, in a household which does not keep chickens, where the average NDVI for in their canine home range is 0; the vegetation coefficient represents the increase in the odds ratio for a dog whose canine home range has an average NDVI of 1, compared to a dog whose canine home range has an average NDVI of 0; the coefficient for living on the coast represents the increase in the odds ratio for a dog living on the coast compared to a dog who does not live on the coast; the coefficient for keeping chickens represents the increase in the odds ratio for a dog whose owners keep chickens compared to a dog whose owners do not keep chickens. We found that an increase in NDVI of 0.1 led to a 1.21-fold increase in the odds of becoming infected with canine visceral leishmaniasis. The vegetation and coast indicator parameters were significant in the model. The indicator of keeping chickens was not significant, and thus odds predictions were carried out for the case where all households do not keep chickens.

An indicator for whether other infected dogs were living in the household was found to correlated with both NDVI and canine VL cases, however, this was not included in modelling since this information was captured by the spatial aspect of the analysis.

Analysis of sandfly data in Camaçari by Mota *et al* [40] found that a large majority of captured sandflies were in beachfront areas, which is an uncommon finding in other areas of Brazil and the rest of the world. They suggest that this may be due to an influx of tourism and urbanisation along the coast in Camaçari. Bahia has undergone rapid urbanisation and an influx of tourism in recent years [40], which has led to an inundation of tourists with low-to-no natural immunity to the disease staying near beaches [6, 13]. Another explanation is that this observation may be due to the type of vegetation present on the Camaçari coast being more attractive to sandflies. Exotic and ornamental plants are often placed in gardens and homes, whereas native plants can grow in less managed areas. This suggests that future studies should also collect more information should thus be collected on the type of vegetation to better understand how this can impact VL infections in dogs. Furthermore, information on the type of vegetation could also be combined with information on the movement of free-roaming domestic canines to better quantify the total exposure to VL.

The inclusion of vegetation as a covariate lead to a decrease in the estimated variance in unexplained residual spatial variation, *σ*^2^, which suggests that the vegetation parameter explained some of the residual spatial variation in the data, and so was beneficial to the model. Both models had a similar scaling parameter, *ϕ*, so the spatial correlation was estimated to persist over a similar range. The model containing vegetation as a covariate had a lower average standard error of the predictions than the model without vegetation as a covariate, thus suggesting that vegetation provided more certainty about predicting the odds of canine VL.

The predicted odds map in Fig 5 highlights the connection between canine VL and both vegetation and coastal areas. The highest odds is found near to the coast in Monte Gordo, where the most vegetation is present compared to the rest of the coastline. The capital city shows the lowest estimated odds, as there is a very small amount of vegetation present. There are more vegetated areas in the south of Abrantes and north of Monte Gordo, however these are more rural than the coastline. Leishmaniasis has undergone rapid urbanisation since the 1980s and is no longer a rural disease in Brazil [41]. Due to the evolution of the transmission cycle, rural areas are no longer as high risk as the urban areas with vegetation present. Nonetheless, the rural areas were given a higher probability of the odds exceeding 1 than the capital city. However, the vegetation coefficient had the most influence on the overall odds in the rural areas of Camaçari, which had the most vegetation present. This indicates that measuring vegetation alone is not sufficient to quantify the risk of canine VL; whether the area is considered urban or rural is also an important factor in determining the risk.

The most commonly used vegetation measure, Normalised Difference Vegetation Index (NDVI), was used to quantify the vegetation in the study area. However, this measure can be significantly affected by the solar incidence angle, time of day, the presence of air moisture, and soil reflectance [42]. The Enhanced Vegetation Index (EVI), a newer method which can correct for the atmospheric issues present in NDVI [42], was also tested, but we did not find an appreciable difference between the two measures for the considered study site. Several newer methods of quantifying vegetation from satellite imagery are also available, such as the Red-Edge Chlorophyll Vegetation Index (RECl), which determines the chlorophyll content in leaves, and the Soil Adjusted Vegetation Index (SAVI), which can correct for soil noise effects in imaging. However, RECl is more useful during the stage of active vegetation development but not suitable for the season of harvesting, and SAVI is most useful at the beginning of the crop production season but less effective during the rest of the year when the majority of the soil of covered by vegetation growth. One of the constraints faced in this study was that, due to large amounts of cloud cover during the study period, it was not possible to use images exclusively from the season of active vegetation development or the beginning on crop production, so neither RECl or SAVI would have significantly improved the vegetation indexing compared to NDVI. Additionally, the Visible Atmospherically Resistant Index (VARI) would be a better measure of the vegetation than NDVI, since it makes use the whole visible spectrum rather than just the red band. Due to this, VARI can significantly reduce the atmospheric effect on findings [43]. However, this measure requires the use of imagery from newer satellites such as Sentinel-2 or Landsat-8, which had not been launched during the study period for this analysis.

Another limitation of our work is that the statistical analysis in this study assumes that dogs were randomly sampled from the population at risk. It was not possible to sample some streets for security reasons. Canines living in informal settlements on the outskirts of the capital city of Camaçari are often at high risk of VL as a result of reduced access to healthcare and poor sanitary conditions [44]. These dogs also tend to come into contact with less vegetated areas than dogs living in rural or coastal areas of Camaçari. However, they were less likely to be sampled than the rest of the canine population due to the presence of criminal gangs. As a result of this, our estimated strength of association between canine VL and vegetation may have been slightly overestimated.

In conclusion, our study finds that the presence of vegetation leads to an increase in VL risk in after accounting for other spatial confounders using a geostatistical model. However, additional information on the type of vegetation and more accurate classification of vegetation at fine spatial scale would allow for a better understanding of how vegetation affects the risk of occurrence of canine VL cases.

## Data Availability

All relevant data are within the paper and its Supporting Information files.

## Supporting information

**S1 Fig**.

**S2 Fig**.

## Acknowledgments

The authors would like to thank the Camaçari Municipal Secretary of Health and the staff of the Camaçari Zoonosis Control Center for their assistance. We especially thank Marcelo Bordoni, Maiara Arruda, Bruna Leite, Miriam Rebouças, Adriana Oliveira de Almeida, Sandra Maria de Souza Passos, and all the municipal health agents for their invaluable assistance with our work in the endemic area. We thank all the dog owners who permitted us to follow their animals in the endemic area. The authors also would like to thank Andrezza Souza for technical and logistics support, Amaro Nunes da Silva for providing assistance during visits to the endemic area and all the students who helped in the execution of this study.

## Author Contributions

**Conceptualization**: Manuela da Silva Solcà, Deborah Bittencourt Mothé Fraga.

**Data Curation**: Freya N. Clark.

**Formal Analysis**: Freya N. Clark, Emanuele Giorgi.

**Funding Acquisition**: Freya N. Clark, Emanuele Giorgi.

**Investigation**: Manuela da Silva Solcà, Deborah Bittencourt Mothé Fraga.

**Methodology**: Freya N. Clark, Manuela da Silva Solcà, Deborah Bittencourt Mothé Fraga, Emanuele Giorgi.

**Project Administration**: Deborah Bittencourt Mothé Fraga, Claudia Ida Brodskyn.

**Resources**: Manuela da Silva Solcà, Deborah Bittencourt Mothé Fraga, Emanuele Giorgi.

**Software**: Freya N. Clark, Emanuele Giorgi.

**Supervision**: Emanuele Giorgi.

**Validation**: Freya N. Clark.

**Visualization**: Freya N. Clark.

**Writing - Original Draft Preparation**: Freya N. Clark.

**Writing - Review & Editing**: Freya N. Clark, Manuela da Silva Solcà, Deborah Bittencourt Mothé Fraga, Emanuele Giorgi.

## Notes

### Competing Interest Statement

The authors have declared no competing interest.

### Funding Statement

Yes

